# Predictors of abnormal Thompson score in term neonates in a tertiary hospital in Zimbabwe

**DOI:** 10.1101/2023.12.06.23299608

**Authors:** N. Khan, E. Mugwagwa, M. Cortina-Borja, E. Catherall, F. Fitzgerald, S. Chimhuya, G. Chimhini, H. Gannon, C. Crehan, M. Mangiza, M. Heys, the Neotree team

## Abstract

**Background:** Neonatal encephalopathy, abnormal neurological function in a baby born at term is a key cause of neonatal death. In the absence of adequate training and brain imaging or monitoring in low-resource settings, clinical risk scores, such as Thompson score, have been useful to predict risk of neonatal encephalopathy. A clearer understanding of the clinical and maternal predictors of abnormal values of Thompson score would be beneficial to identify term neonates with suspected neonatal encephalopathy.

**Methods:** A scoping review of the literature identified a set of *a priori* neonatal and maternal variables associated with neonatal encephalopathy in low-resource settings. Next, a prospective study of all neonates born at term admitted to Sally Mugabe Central Hospital in Zimbabwe between October 2020 and December 2022 (*n*=6,054) was conducted. A predictive statistical model for abnormal (>10) Thompson score (range 0-22) was developed.

**Results:** In total 45 articles were identified from three databases and 10 articles were selected. 45 candidate predictors were identified −36 from the available literature and 9 from clinical data and experience. 4.06% (*n*=246) of neonates had an abnormal Thompson score of 10 or more on admission and 90.65% (*n*=223) of these neonates had an Apgar score less than 7 at 5 mins (*p*<0.001). 24 possible predictors were selected as the most important of which nine factors were identified as the most useful in predicting which neonates are at risk of abnormal Thompson score. These predictors and their adjusted odds ratios are: low Apgar score at 5min (OR= 0.46, 95%CI=(0.42, 0.51)), low neonatal heart rate at admission (OR=0.977, 95%CI=(0.97, 0.985)), temperature lower than 36.5°C (OR=1.64, 95%CI=(1.18, 2.28)), abnormal head shape (OR=2.12, 95%CI=(1.51, 2.97)), resuscitation received (OR=3.95, 95%CI=(1.69, 11.01)), neonatal encephalopathy as an admission reason (OR= 2.47, 95%CI=(1.37, 4.32)), risk factors of sepsis other than premature rupture of membrane and offensive liquor (OR=2.04, 95%CI=(1.1, 3.67)), respiratory distress as an admission reason (OR=2.48, 95%CI=(1.59, 3.96)), and other admission reasons (OR=1.81, 95% CI=(1.12, 2.97)). The main admission reasons in ‘Other’ category include low birthweight, meconium aspiration and hypoxic ischaemic encephalopathy and congenital abnormality.

**Conclusion:** In resource-poor settings where it may be not possible to clinically assess all admitted neonates, those with the identified risk factors should be prioritised for a Thompson score assessment. Local clinical guidelines should incorporate these factors into the clinical management of at-risk neonates and assess their impact on clinical care and neonatal outcomes.

## Introduction

Progress has been made globally in reducing under-5 mortality; however, mortality in newborns (below 28 days of age) remains disproportionately high, accounting for nearly half of under-5 deaths (World Health Organization, 2020). Globally 2.4 million neonates die within the first 28 days of life. Of these deaths, 90% occur in low-to-middle income countries, with the highest neonatal mortality rates occurring in sub-Saharan Africa (Liang et al., 2018). The three commonest causes of mortality are infection, prematurity and neonatal encephalopathy (NE) (Ou et al., 2022).

NE is typified by abnormal neurological function in the first few days of life in a baby born at term (gestational age of 37 weeks or more). It can be defined as a neonatal neurological syndrome characterised by depressed levels of consciousness, seizures, abnormal tone, respiratory depression and impaired feeding (Volpe, 2012). Among others, NE can be caused by perinatal asphyxia, metabolic disorders, perinatal infections and placenta abnormalities (Aslam et al., 2019). Hypoxic Ischaemic Encephalopathy (HIE) is a condition characterised by the presence of NE with evidence of hypoxic-ischaemia as the cause (Kurinczuk et al., 2010). In addition to causing an estimated one million deaths worldwide every year, NE is a cause of significant life-long disability (Tann et al., 2018). However, clinical diagnosis and management is difficult in most low-resource settings where the majority of these neonates are born.

In high-resource settings, the diagnosis of NE and identification of its underlying risk factors are determined by clinical assessment and investigation of the mother and baby including cord blood arterial blood gases, neuroimaging procedures such as continuous amplified electroencephalogram (EEG) and magnetic resonance imaging (MRI) (Vannucci, 2000). Continuous amplified EEG and MRI are rarely available in low-resource settings where diagnosis is made on clinical features alone. Hence it is often challenging to diagnose NE with certainty in such settings, or differentiate between NE caused by perinatal asphyxia or other causes.

Clinical prediction scores combining clinical features with EEG result have been developed to categorise severity of NE and predict neurodevelopmental sequelae in neonates with a well-defined episode of foetal distress, such as the Sarnat and Sarnat score (Sarnat & Sarnat, 1976). The Sarnat and Sarnat score has two key limitations, namely the reliance on EEG and a need to incorporate a well-defined episode of foetal distress - neither of which are routinely available in low-resource settings due to lack of monitoring during labour. To address these gaps, Thompson et al. (1997) developed a simplified score incorporating findings on clinical examination alone. The Thompson score relies on nine clinical features and assigns a score of 0 to 2 or 3 per feature to characterise peak severity of encephalopathy in term neonates (range 0-22). Infants with a score of 0 are considered normal, those with a score between 1 and 10 are considered to have a diagnosis of mild NE; a score 11-14 is considered indicative of moderate NE and a score of 15-22 as severe NE. The Thompson score is mainly reported in the literature to examine and characterise NE in cohorts that are already suspected of perinatal asphyxia and NE.

A high value of Thompson score within the first seven days of life is a sensitive predictor of abnormal neurodevelopmental outcome (Thorson et al., 2016; Weeke et al., 2017; Mendler et al., 2018). However, criteria for measuring an infant’s Thomspon score, namely: “*if clinical signs of hypoxic–ischemic encephalopathy developed after birth*” are unclear and difficult to operationalise in neonatal units where clinical staff may not be sufficiently skilled in identifying clinical signs of NE (Thompson et al., 1997). Therefore, the underlying reasons for suspecting NE and subsequently wanting to measure the Thompson score are neither clearly stated nor uniform across different studies (Badawi et al., 1998; Biselele et al., 2014). Nevertheless, these studies provide valuable evidence for the various maternal and neonatal factors that may trigger clinicians to use the Thompson score. For example, Badawi et al. (1998), included maternal criteria such as severe pre-eclampsia and moderate-severe vaginal bleeding during pregnancy in increasing risk of NE and thus a reason to calculate a Thompson score. Further work is needed to clarify the prognostic value of the Thompson score in the first few hours of life or at admission to a neonatal unit. This will help the healthcare providers identify the neonates at risk of NE on admission and therefore should be prioritised for examination and assessment.

As part of a wider programme of work in Malawi and Zimbabwe a digital health intervention called Neotree has been developed to improve neonatal care and outcomes (Heys et al., 2022, Khan et al 2022, Crehan et al., 2019, Gannon et al., 2021). Neotree provides data capture, clinical decision support and education in newborn care, with accompanying process for data collection, curation, visualisation, and export (Khan et al., 2022). It has been embedded as part of daily care at two sites in Zimbabwe (Sally Mugabe Central Hospital and Chinhoyi Provincial hospital) and one site in Malawi (Kamuzu Central Hospital). A clinical diagnostic algorithm for NE is being developed within this wider programme of work. At first, international and national evidence-based guidelines describing risk factors for NE were digitalised. Next, a Delphi review was conducted by a panel of international experts in newborn care, who reviewed the NE algorithm proposed by the Neotree team and concluded that there was a lack of evidence for these risk factors to be used for NE diagnosis (Evans et al., 2021). The use of validated encephalopathy scores such as the Thompson score was recommended instead (Biselele et al., 2014) to guide the diagnosis and clinical management of NE. However, due to a shortage of skilled healthcare professionals in low-resource settings, it is not always feasible to conduct a Thompson score assessment on all term neonates admitted to newborn care units (Crehan et al. 2019). Furthermore, it remains unclear which neonates should be prioritised for a detailed neurological and Thompson score assessment (Mrelashvili et al., 2020).

The goal of this study is to characterise neonates at risk of NE born in low-resource settings where routine access to blood tests or neurological investigations can be scarce. Our aim is to develop a statistical predictive model for an abnormal Thompson score values that can be incorporated into clinical pathways for newborn care to enable healthcare providers to consistently and explicitly decide which infants to screen for NE using the Thompson Score. *A priori* variables were identified to develop a predictive model for abnormal Thompson score values. This helps us to understand the role of neonatal and maternal predictors which identify neonates in need of measuring their Thompson score. A scoping review to determine a priori predictors of abnormal Thompson score was conducted, and. routine data collected from a neonatal intensive care unit of a tertiary hospital in Zimbabwe were analysed.

## Methods

### Step 1: Scoping review

To identify maternal and neonatal clinical and demographic factors that might be associated with abnormal Thompson score and thus a high likelihood of diagnosis of NE, three search engines were used to find articles published up to 31 July 2022: Medline (PubMed), Scopus and Web of Science. The following PICO (Population, Intervention, Comparison, Outcome) terms were used for the literature search and the exact search strings are included in the supplementary material:

- Population: Neonates born in low resource settings
- Intervention/exposure: Any maternal, perinatal and neonatal factors predictive of NE
- Comparison: None
- Outcome: NE cases with abnormal Thompson score, Sarnat score or Hypoxic ischaemic encephalopathy score.

Titles and abstracts were evaluated for eligibility. Inclusion criteria for full-texts included: a) neonates; b) with a diagnosis of (i) hypoxic ischaemic encephalopathy or NE or (ii) birth asphyxia or (iii) perinatal asphyxia; c) assessed using Thompson score, Sarnat score or Hypoxic ischaemic encephalopathy score; d) studies that also included maternal factors; and e) studies in low-income, low- and middle-income countries or low-resourced settings. Exclusion criteria included: a) animal studies; b) studies not published in English; c) use of scoring systems other than Thompson score, Sarnat score or Hypoxic ischaemic encephalopathy score; d) clinical trials; e) not including risk factors or predictors of NE.

### Step 2: Modelling of risk factors associated with abnormal Thompson score

#### Study setting and population

All neonates born at term (gestation age ≥ 37 weeks) who were admitted in the neonatal unit of Sally Mugabe Central Hospital in Zimbabwe and were assessed for Thompson score were included in the study. Data were collected as per usual clinical care at the bedside using Neotree and pseudonymised prior to analysis (Heys et al., 2022). 1^st^ October 2020 to 31^st^ December 2022 was selected as the study period as Thompson scores were regularly taken for all term neonates during this time. The hospital has a 100-bed neonatal unit and on average 225 monthly admissions. Available equipment include oxygen and non-invasive ventilation but there is no access to brain imaging, electroencephalogram, or cord blood samples to clinically assess NE in neonates (Gannon et al., 2022). Neonatal mortality rate for the study period was 112 per 1000 admissions (95%CI = (103, 121)).

#### Main outcome measure

Thompson et al. (1997) showed that in normothermic infants, a score >10 in the first 7 days of life can predict abnormal neurodevelopmental outcomes with 100% sensitivity and 61% specificity.

Therefore, the main outcome is Thompson score >10 at admission.

#### Predictive variables

The predictive variables were derived from the scoping review in step 1 and from existing national and international guidelines that were explored in a previous Delphi study (Evans et al., 2021). This mapped list of candidate predictors was reviewed by a group of six neonatal experts working in Zimbabwe and Malawi. The experts were asked to evaluate the initial list of predictors and add any other predictors they thought were missing, based on their clinical experience. 6 potential predictors were identified from this process. Neotree data dictionary was reviewed to identify which predictors are included in the data collected through Neotree and the final set of predictive variables was created.

#### Data Analysis

Missing data in categorical variables were coded as a ‘missing’ category. Numeric predictive variables were assessed to ascertain whether data were missing at random or not at random and imputed using conditional medians. Generalised linear modelling was used to determine the effect that the selected *a priori* variables had on the probability that a neonate had a Thompson score >10. Association between categorical variables was analysed using χ^2^ test. To prevent overfitting the dataset was limited to only those input variables that were most predictive for the outcome variable. Logistic regression models were fitted for the final set of variables and Akaike’s Information Criterion (AIC) was used as a criterion of goodness-of-fit.

All analyses were performed in the R statistical language (version 4.1.2) on R Studio (R Core Team, 2021).

## Results

### Scoping review

In total 45 papers were identified from three databases and three were excluded for not meeting the language criteria. 42 articles were assessed for eligibility and 32 were excluded for reasons included in Figure 1. This resulted in 10 papers that described Thompson score or Sarnat grading or HIE score in neonates in low-and-middle income countries with known or suspected diagnosis of NE (or HIE/BA/PA etc). These papers included description of maternal, perinatal or neonatal factors that might be associated with a diagnosis of NE/HIE/BA/PA.

**Figure 1:**
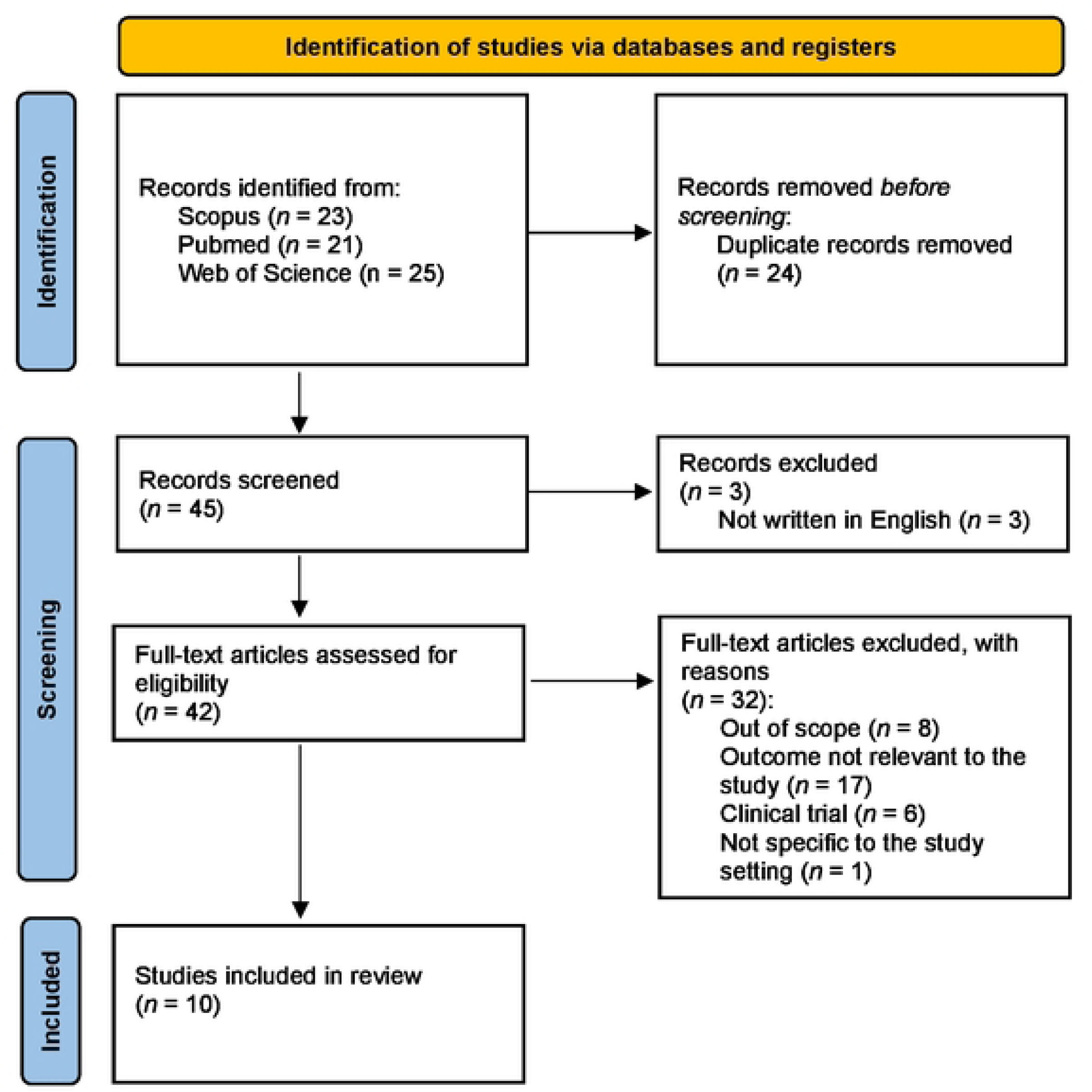
PRISMA flow diagram.

Table 1 includes the details of the studies selected, including study type, sample size, study setting, inclusion criteria, and the scoring system used. Out of 10 identified studies, three studies were prospective cross-sectional (Bhagwani et al., 2016; Biselele et al., 2013; Sadhanidar et al., 2022), two were prospective observational (Biselele et al., 2014; Doreswamy & Ramakrishnegowda, 2021), two were prospective cohort studies (Horn et al., 2013; Mwakyusa et al., 2009) and three were retrospective (<u>F</u>utrakul et al., 2006; Shah et al., 2005; Stofberg et al., 2020). These studies were conducted in India (*n*=3), Nepal (*n*=1), Thailand (*n*=1), Democratic Republic of Congo (*n*=2), South Africa (*n*=2), and Tanzania (*n*=1). Sample sizes ranged from 20 to 145 and all described term neonates admitted with a diagnosis of perinatal asphyxia. Low Apgar score (<6 or ≤7) was included as an inclusion criterion to determine perinatal asphyxia in seven out of 10 of the studies and six studies described the use of Thompson score to determine NE.

**Table 1:** Studies selected from the scoping review.

| Study | Sample | Study design | Setting | Inclusion criteria | Scoring system used |
| --- | --- | --- | --- | --- | --- |
| Bhagwani et al. (2016) | 145 | Prospective, cross-sectional | Hindu Rao Hospital, New Delhi, India | Full-term neonates with low-Apgar score, i.e., 1 min score of $\leq 7$ | Thompson score |
| Biselele et al. (2013) | 50 | Prospective, cross-sectional | University Hospital of Kinshasa, Congo | Term and near-term newborn | Sarnat grading and Thompson score |
| Biselele et al. (2014) | 20 | Prospective, observational | University Hospital of Kinshasa, Congo | Term and near-term newborn, Apgar <6 after 5 min, Resuscitation until 10 min after birth, pH <7 and base excess more than 16 on umbilical cord blood or any blood sample (arterial, venous or capillary) within 60 min of birth, Thompson score > 5 one hour after birth. | Thompson score |
| Doreswamy and Ramakrishnegowda (2021) | 55 | Prospective observational | India | Neonates > 35 weeks of gestation with birth weight $\geq 1800$ g and requiring resuscitation at birth | Thompson score |
| Futrakul et al. (2006) | 84 newborns with perinatal asphyxia | Retrospective | King Chulalongkorn Memorial Hospital, Thailand | Term neonates ($\geq 36$) weeks of gestation with Apgar score < 6 at 1 minute | Modified Sarnat-Sarnat Score |
| Horn et al. (2013) | 41 | Prospective cohort study | Three hospitals in Cape Town, South Africa | Gestation week $\geq 36$ and birth weight $\geq 2000$ g, with signs of encephalopathy<br>after age 10<br>minutes but before<br>age 5 hours. 10-min Apgar score<br><7 or continued<br>respiratory<br>support at age 10<br>min. | Thompson score |
| Mwakyusa et al.<br>(2009) | 140 | Prospective<br>cohort study | Muhimbili<br>National<br>Hospital, Dar-<br>es-Salaam,<br>Tanzania | Full-term neonates<br>with low-Apgar<br>score (i.e. a 5 min<br>score $\leq 7$) or post-<br>asphyxial<br>symptoms<br>admitted within 24<br>h of delivery or<br>with symptoms<br>and signs of HIE. | HIE score<br>(modified<br>Sarnat<br>scoring<br>system) |
| Sadhanidar et al.<br>(2022) | 50 | Prospective<br>cross-sectional | A tertiary care<br>government<br>hospital in<br>Guwahati,<br>Assam, India | All<br>haemodynamically<br>stable newborns<br>irrespective of the<br>gestational age<br>sustaining<br>perinatal asphyxia<br>with clinical diagnosis of HII were included. | Sarnat and<br>Sarnat<br>Staging<br>system |
| Shah et al. (2005) | 50 | Retrospective | B.P. Koirala Institute of Health Sciences, Dharan, Nepal | Gestational age $\geq 37$ weeks with Apgar score $\leq 6$ at 5 minutes | Sarnat and Sarnat Staging system |
| Stofberg et al. (2020) | 29 | Retrospective | A district hospital in Cape Town, South Africa | Gestational age $\geq 36$ weeks, birth weight $\geq 1.8$kg, a sentinel event in the perinatal course, Apgar score $\leq 7$ at 5min, prolonged resuscitation needed at birth (more than 10 min), proven acidosis within the first hour or life, and other clinical features | aEEG and Thompson score |

Biselele et al. (2014) studied the evolution of Thompson score over the first 6 hours of life and found that the scores change over this time. In a study to understand the clinical profile of asphyxiated newborns, Shah et al. (2005) found that common presentations of HIE had reflexes, seizures, lethargy, and pupillary abnormalities. Subtle seizure, meconium-stained liquor, post-term, and lack of antenatal check-up were reported as key risk factors by Bhagwani et al. (2016). Similar findings were also reported by Futrakul et al. (2006). The authors retrospectively modelled the risk factors of HIE and found inappropriate antenatal care, post-term gestation, vacuum extraction, male gender, prolapsed cord, and Apgar scores at 1 and 5-minute to be significant risk factors of HIE. High maternal age (median 31 years) and a high incidence of pre-eclampsia were found in the study sample of Biselele et al. (2013). The authors compared Thompson score and Sarnat score and found correlation between the two scores, as well as with mortality and individual scoring system. In contrast, Mwakuysa et al. (2008) reported developmental outcome at 6 months of age of 140 neonates diagnosed with birth asphyxia and found that higher Thompson scores were associated with increased risk of death, seizures and poor neurological outcome. Horn et al. (2013) found that Thompson score could be used to predict death or a persistently severely abnormal amplitude-integrated electroencephalogram (aEEG) at 48-hour. The authors suggested that a score ≥16 could be used to identify neonates who will have a poor outcome despite cooling. Stofberg et al. (2020) reported similar findings where neonates with Thompson score ≥12 were associated with abnormal aEEGs, indicative of HIE. Non-operative deliveries, lack of a doctor at the time of delivery and neonatal chest compressions were also associated with abnormal aEEGs. Doreswamy and Ramakrishnegowda (2021) conducted a study to validate Prediction of Encephalopathy in Perinatal Asphyxia (PEPA) score by Holdout method where Thompson score between 3 and 5-hour of life was used to determine post-test probability of developing encephalopathy. The authors found that PEPA score had a higher sensitivity than National Institute of Child Health and Human Development (NICHD) criteria for prediction of HIE in asphyxiated neonates.

From these studies a list of 36 factors that might be associated with an abnormal Thompson score was compiled (Appendix A). Combining these with the nine predictors identified from clinician consultation and Neotree data, in total 45 potential predictors were identified. 15 of these predictors were either already included as part of the Thompson score or were not currently collected as part of routine data capture via Neotree, leaving 29 potential factors to consider. 11 predictive variables were collapsed into five variables (A and B), leaving 24 candidate measures (Appendix A).

### Modelling

#### Study population

There were 9,445 newborns admitted during the study period of October 2020 to December 2022, and 6,109 term neonates meeting the inclusion criteria. Neonatal mortality rate among term neonates with a recorded outcome was 112 per 1000 admissions (95%CI = (103, 121)).

The Thompson score outcome was missing in 55 (0.9%) and were excluded given the proportion is very small, resulting in a total sample size of 6,054 neonates. Of the 24 candidate predictors, 11 had missing values. For foetal heart rate (0.11%), maternal age (0.33%), and temperature at admission (0.69%) they were imputed using medians. For birthweight (0.26%) and Apgar score (5.2%) they were imputed as medians conditional on the gestational age and Thompson score values, respectively. For categorical predictors, namely resuscitation (3.15%), meconium-stained liquor passed (9.78%), maternal HIV infection (10.3%), meconium-stained umbilicus (11.1%), place of birth (11.7%), and duration of labour (16.09%) a “missing” category was included (Table 2).

**Table 2:**
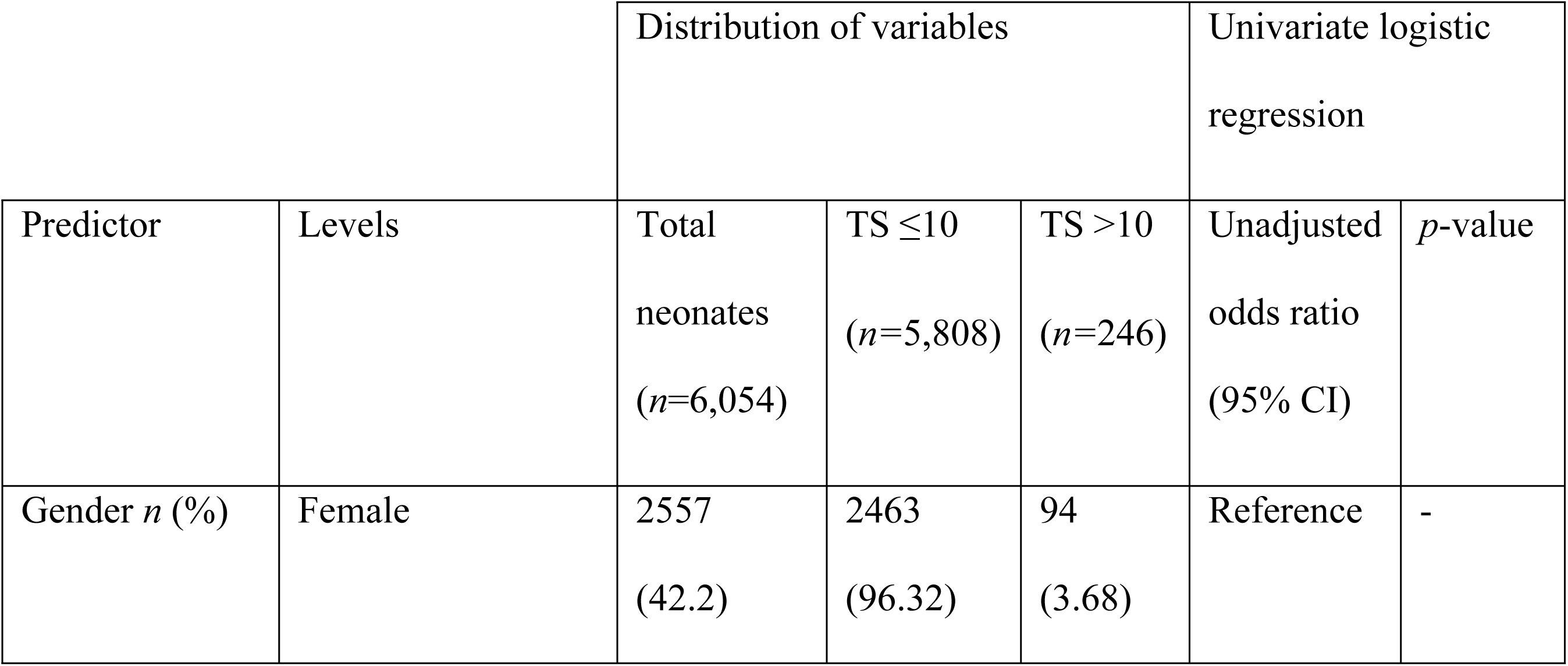
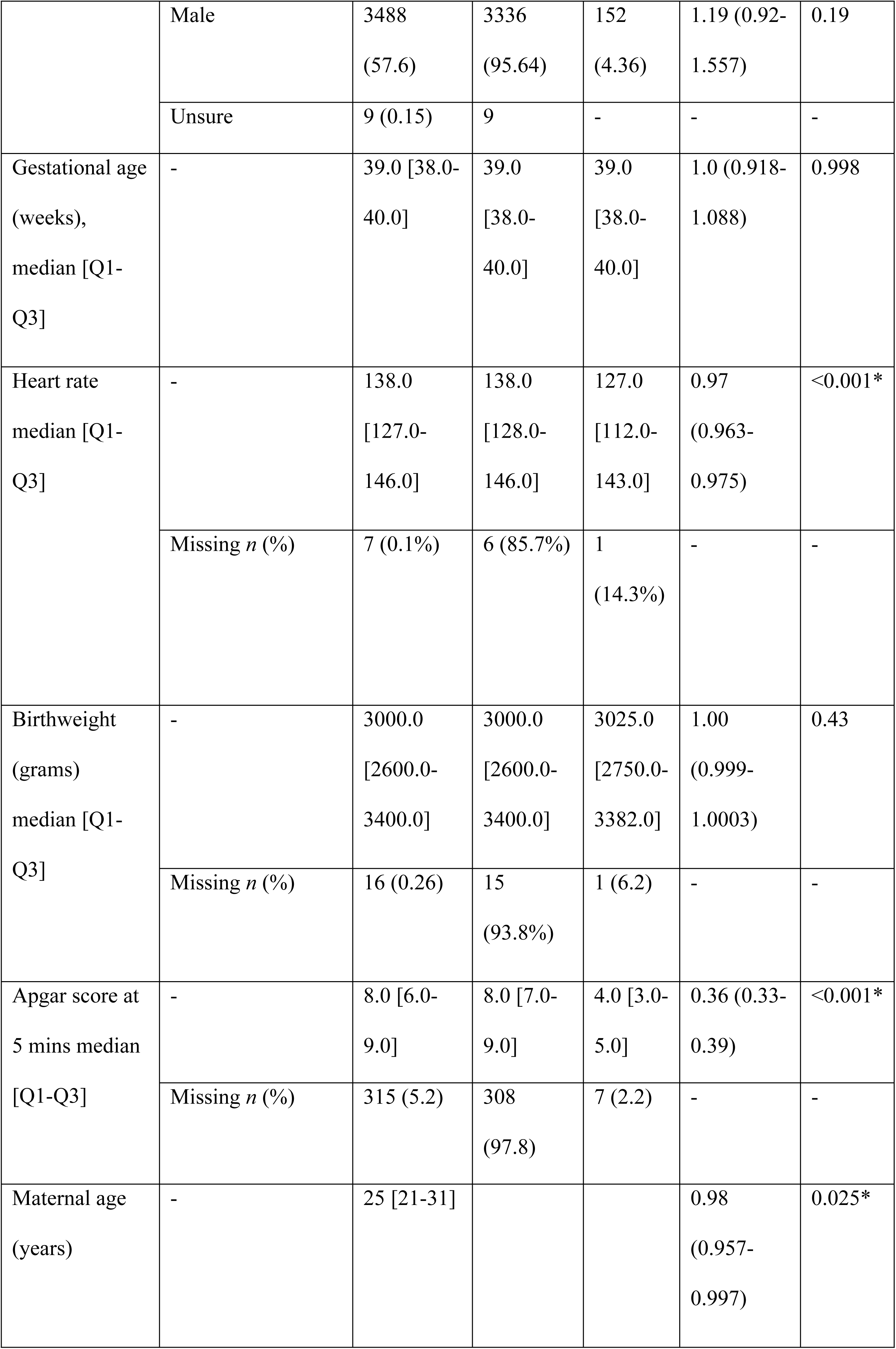
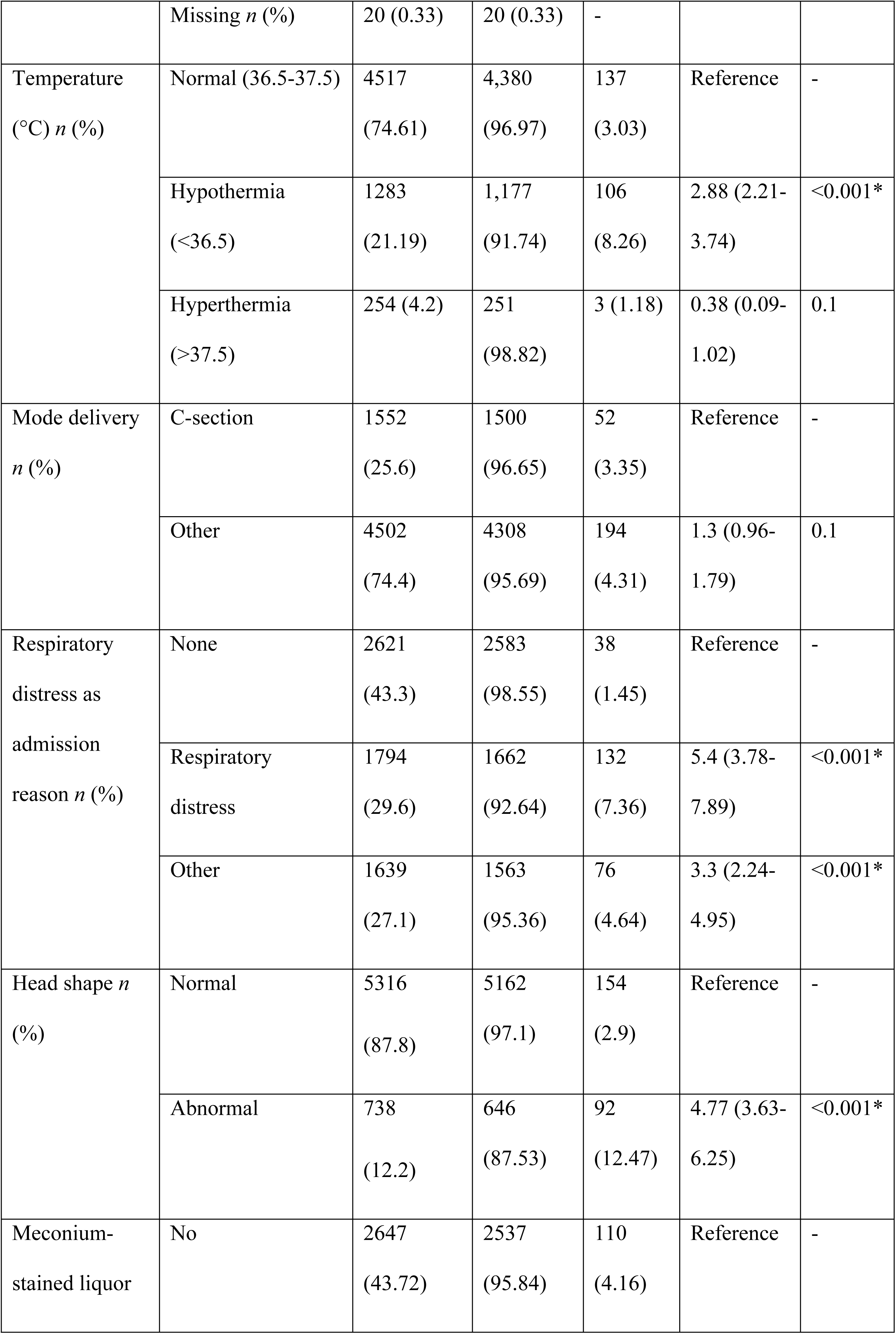
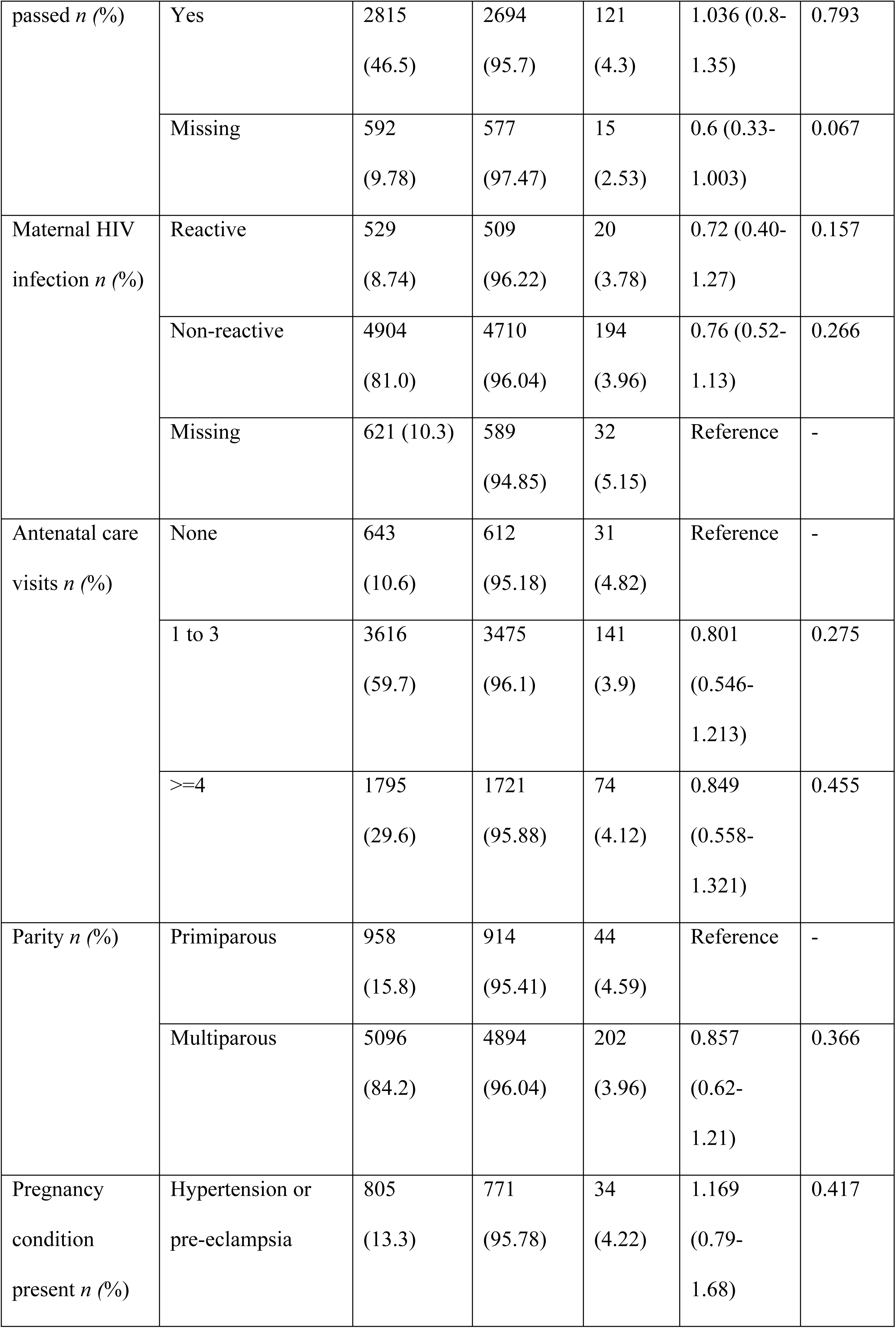
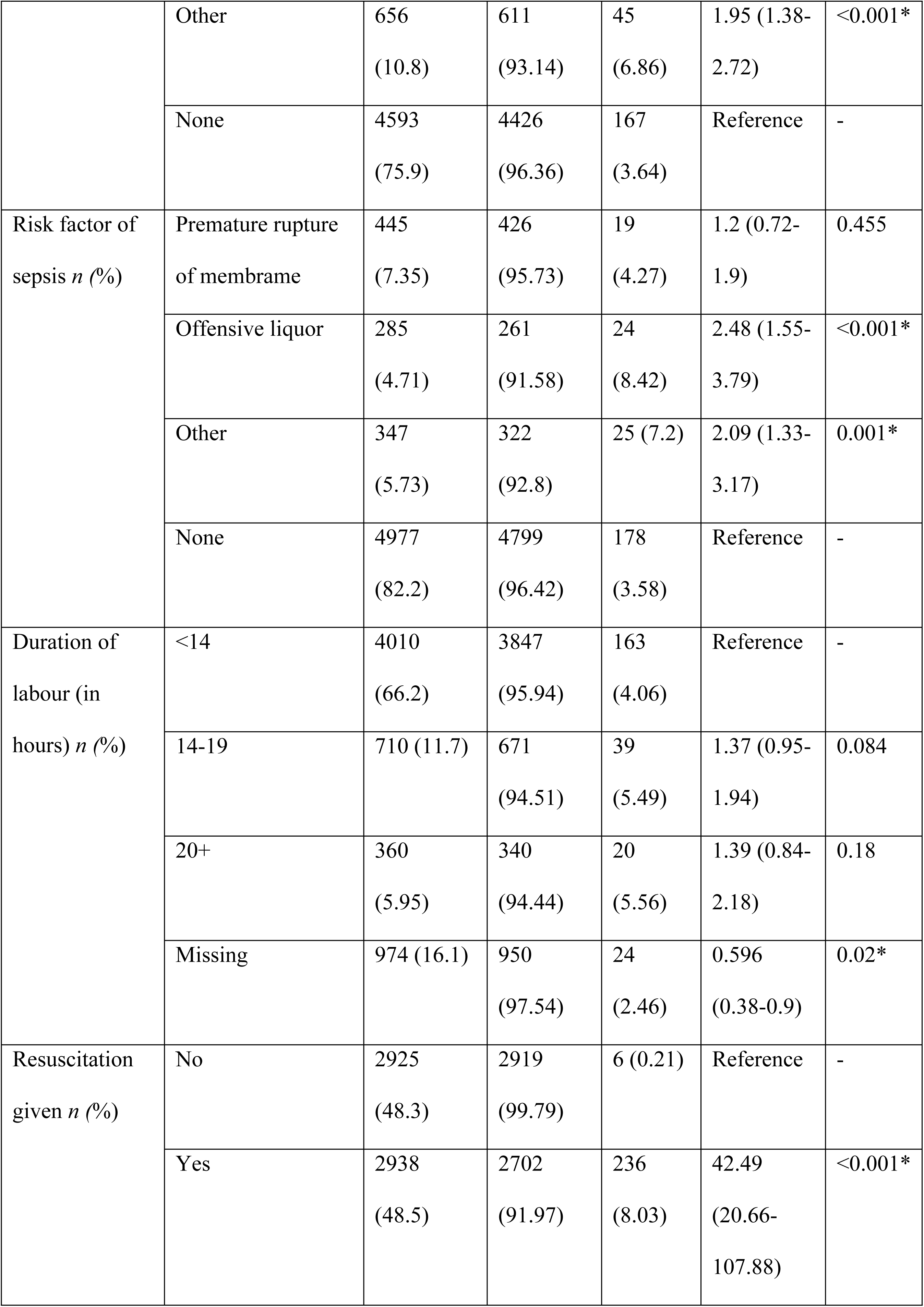
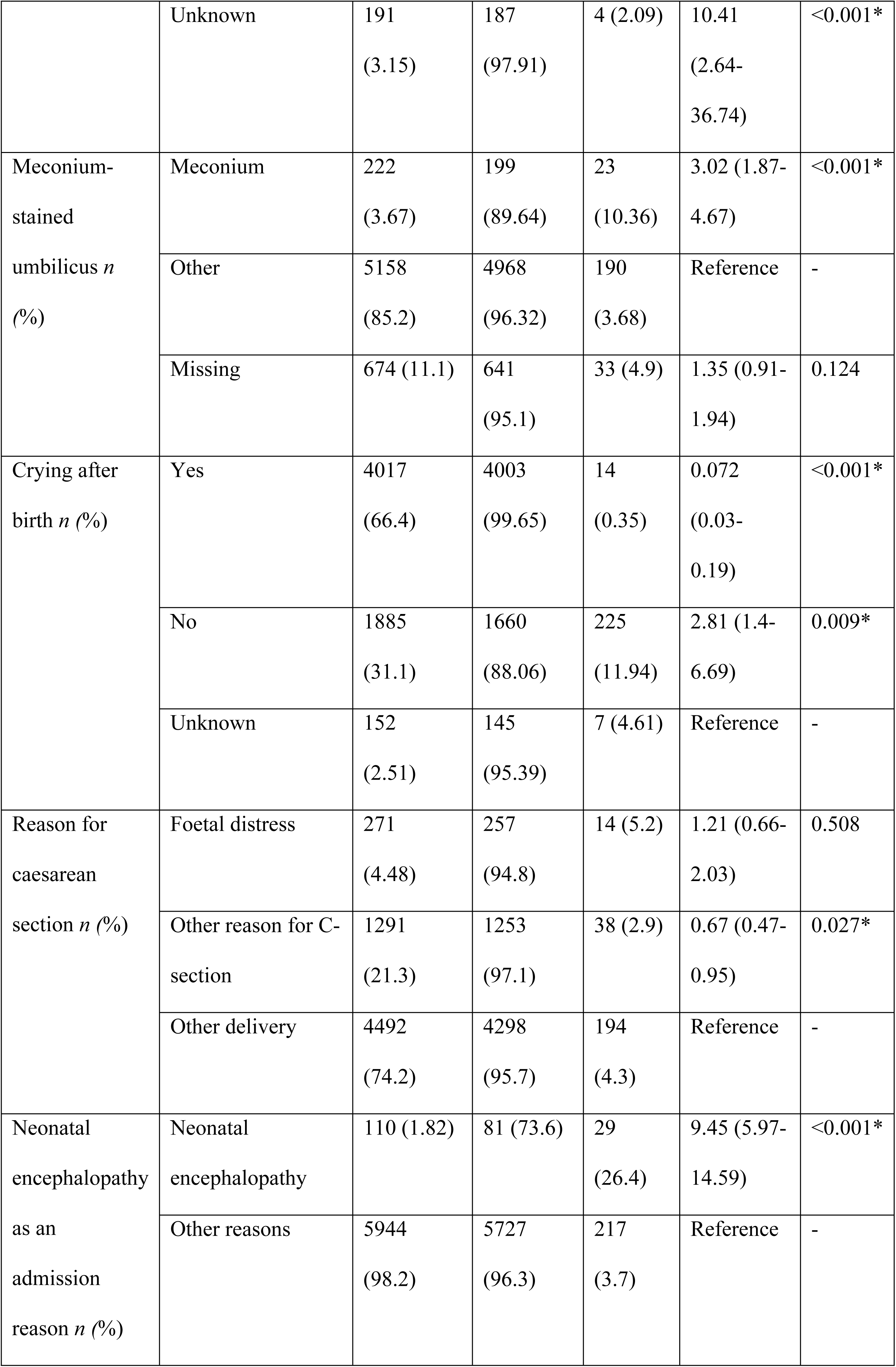
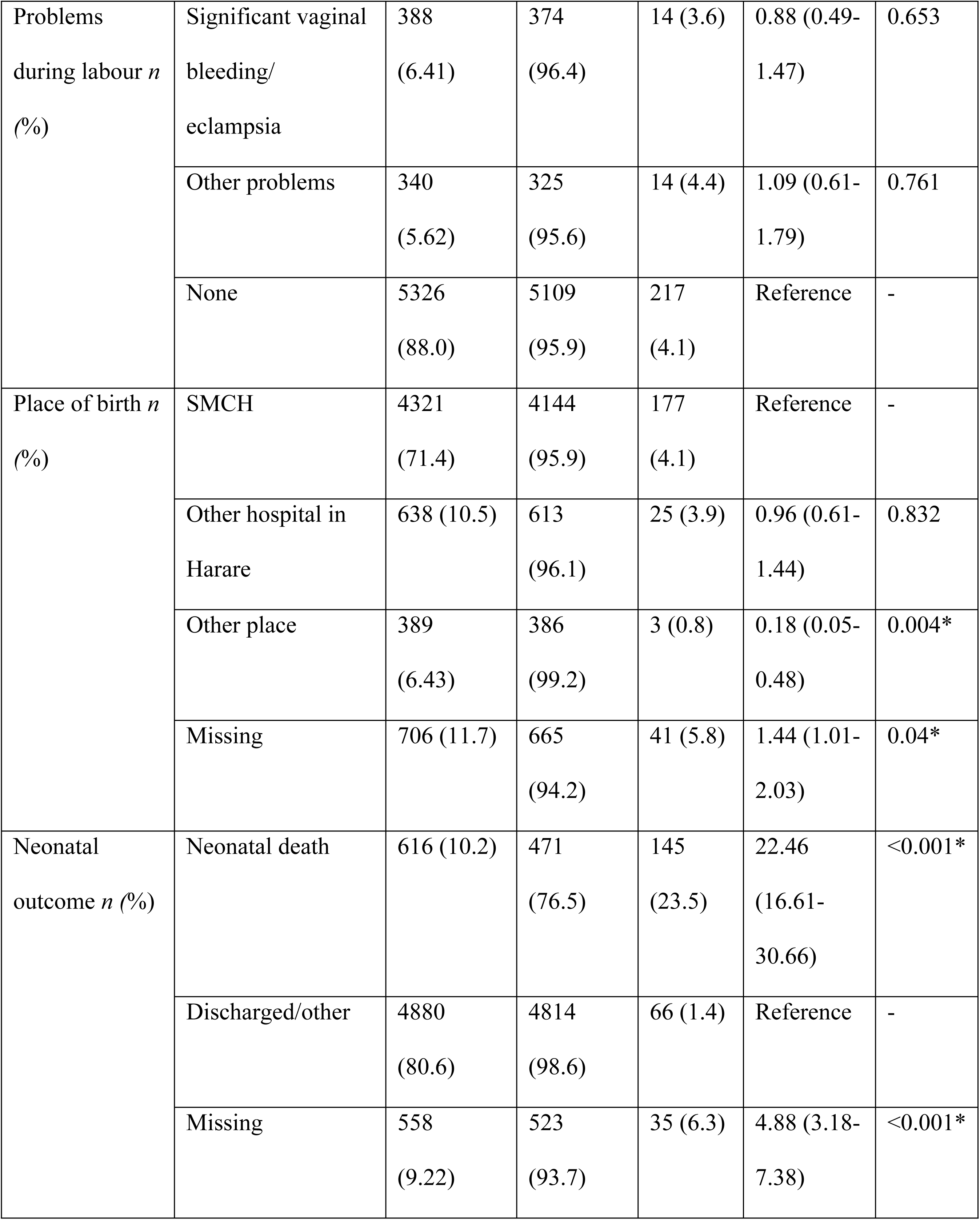
Demographic characteristics of the study sample (*n*=6,054)

Of these 6,054 neonates, 57.6% were male and 70.7% were born via spontaneous vaginal delivery. 558 (9.22%) neonates did not have any outcomes recorded. Among the rest 5,496, 88.8% of newborns survived to discharge, giving an overall case fatality rate of 112 per 1000 admitted term neonates (CI 103.4-121.3). Mean maternal age was 25 years (SD 7). Mean Thompson score was 1.996, Median Thompson score was zero (IQR 0-2). 246 (4.06%) of neonates had a Thompson score of greater than 10 equating to a rate of 40 per 1000 admitted neonates. 511 (8.4%) of neonates had a Thompson score of greater than 7.

For the ‘Total neonates’ column in table 2, counts and percentages are shown out of 6,054. For the ‘TS≤10’ and ‘TS>10’ columns, within the group distribution of counts and percentages are shown. For example, in the Gender variable, 2557 out of 6054 (42.2%) of total newborns were female. Amongst these 2557 newborns with a Thompson score, 2463 (96.32%) had TS≤10 and 94 (3.68%) had TS>10. Compared to those with TS≤10, neonates with TS>10 had significantly lower median Apgar score at 5-minute. Those with TS>10 had 22 times higher odds of neonatal death compared to those with TS≤10.

#### Logistic regression analysis and predictive modelling

Univariable logistic regression models were fitted to test the research hypothesis regarding the relationship between the selected candidate predictors and Thompson score greater than 10. 15 out of 24 of the candidate predictors were significantly associated (Table 2).

Although ‘Other conditions’ for pregnancy conditions variable showed significant association (*p*-value <0.001), it was excluded from multiple logistic regression modelling as it alone includes multiple and ‘unknown’ pregnancy conditions. Thus, it may not be effective in identifying which of those pregnancy conditions is most predictive of an abnormal Thompson score. Similarly, ‘Other place’ in the place of birth variable was significantly associated (*p*=0.004) but has a very small number of cases (*n*=3) and missing place of birth was also significant (*p*=0.04). Due to uncertainty in defining the location for Thompson score greater than 10 this variable was excluded from the final model. For duration of labour variable, only the ‘missing’ category was significant (*p*=0.02) and was excluded for the same reason.

The remaining 12 candidate predictors that were used in multiple logistic regression analysis were: 1. neonatal heart rate (beats/min); 2. Apgar score at 5 minutes; 3. temperature at admission; 4. NE as a reason for admission; 5. respiratory distress as admission reason; 6. head shape; 7. reason for caesarean section (C-section); 8. maternal age (years); 9. crying immediately after birth; 10. duration of resuscitation (minutes); 11. meconium-stained umbilicus and 12. risk factors of sepsis.

Multivariable logistic regression models were fitted for the final dataset of 6054 neonates to test the null hypothesis that there was no relationship between these 12 predictor variables selected from the univariable analysis and a Thompson score >10. Stepwise regression was used for analysis with LRT was used to select the final model. Table 3 shows the regression coefficients and odds ratios for this model.

**Table 3:** Regression coefficients and odds ratios for the final model (*n*=6054)

| Predictor | Estimate | Standard error | Odds ratio | 95% CI | *p*-value |
| --- | --- | --- | --- | --- | --- |
| Intercept | 2.391 | 0.837 | - | - | - |
| Heart rate | -0.023 | 0.004 | 0.977 | 0.97 - 0.985 | <0.001 |
| Apgar score | -0.768 | 0.053 | 0.464 | 0.418 - 0.514 | <0.001 |
| Temperature < 36.5 | 0.496 | 0.168 | 1.642 | 1.178 - 2.278 | 0.003 |
| Abnormal head shape | 0.751 | 0.172 | 2.119 | 1.508 - 2.965 | <0.001 |
| Neonatal encephalopathy as a reason for admission | 0.905 | 0.29 | 2.473 | 1.385 - 4.32 | 0.002 |
| Resuscitation received | 1.375 | 0.471 | 3.954 | 1.69 - 11.008 | 0.004 |
| Respiratory distress as an admission reason | 0.909 | 0.232 | 2.483 | 1.593 - 3.963 | <0.001 |
| Other admission reasons | 0.594 | 0.248 | 1.811 | 1.122 - 2.973 | 0.017 |
| Other risk factors of sepsis | 0.711 | 0.307 | 2.035 | 1.096 - 3.665 | 0.02 |

Abnormal neonatal heart rate on admission (<100 bpm) and lower Apgar score at 5-minutes were associated with higher odds of Thompson score >10. The rest of the predictors were associated with increased odds of a Thompson score greater than 10 with the highest being receiving resuscitation. Hypothermia at admission (Temperature < 36.5) was found to be associated with increased odds of NE.

## Discussion

To our knowledge, this is the first large-sample prospective study in a low-resource setting exploring associations between abnormal Thompson score and thus risk of NE on admission of term infants (gestation age > 36). NE is a clinical diagnosis in low-resource settings where investigations such as EEG, cranial imaging and umbilical cord gases are typically unavailable. Thompson score is a key scoring system used to identify at-risk neonates. However, it may be unclear which neonates should be assessed for clinical signs of NE and thus necessitate the measurement of Thompson score. This study aimed to identify key risk factors of neonates with abnormal Thompson score (>10) based on data collected using Neotree from 6,054 neonates admitted at a tertiary-level hospital in Zimbabwe. The prevalence of abnormal Thompson score (as a proxy of diagnosis of NE) in this population of term neonates was 40 per 1000 with 58.9% (145 out of 246) of these neonates dying. It is not possible to compare this with prevalence estimates per 1000 live births. These estimates are lower than a recent study in Nigeria (Ezenwa et al., 2021) reporting prevalence of 71 per 1000 admissions with 25.3% respectively where NE was defined in a term infant as the presence of encephalopathy or altered consciousness and multi-organ failure with a positive history of delayed cry at birth or required prolonged resuscitation at birth in addition to the presence of any of the neurological features as contained in the Sarnat and Sarnat classification.

Initially, 43 predictors were identified based on literature review and consultations with clinicians in Zimbabwe. Of these, 24 were selected to be most important. Nine risk factors were identified using logistic regression, the presence of which might prompt healthcare providers to undertake a neurological examination and calculate a Thompson score. These are the most predictive covariates for determining a Thompson score of greater than 10: Apgar score at 5-minute, neonatal heart rate on admission, hypothermia at admission, resuscitation, abnormal head shape, NE and respiratory distress as admission reasons along with others; and maternal risk factors of sepsis other than premature rupture of membrane and offensive liquor.

Lower Apgar score was associated with higher odds of abnormal Thomson score, which was in line with the studies from scoping review where all studies used low Apgar score as an inclusion criterion. In a similar study conducted by Futrakul et al. (2006) the authors found significant association of NE with Apgar score at 5-minute. NE as admission reason increased the odds of an abnormal Thompson score by twice. Although maternal pre-eclampsia was one of the prenatal diagnoses reported by Biselele et al. (2013) and Biselele et al. (2014), it was not significantly associated in this study population. Receiving resuscitation at birth significantly increased the odds of abnormal Thompson score. This is plausible as previous studies used resuscitation until 10-minutes after birth as an inclusion characteristic for perinatal asphyxia besides low Apgar score (Biselele et al., 2014). Low neonatal heart rate on admission suggests that neonates who may be in extremis are being transferred to the neonatal unit reflecting severity of insult around the time of birth. Although some studies reported inappropriate or absence of antenatal care as a maternal risk factor (Futrakul et al., 2006; Bhagwani et al., 2016), this risk factor was not strongly associated in this study population. This study identified that maternal risk factor of sepsis, other than premature rupture of membrane and offensive liquor can increase the odds of Thompson score greater than 10.

A key strength of this study is that the *a priori* variables used in predictive modelling were assembled using a combination of evidence from the literature, routine health records and clinically driven reasoning. In this way, the study is grounded in the lived experiences of clinicians, data collection systems and healthcare providers in low-resource settings who often have limited data and resources needed save the lives of infants. Compared to previous studies that modelled risk factors of NE (Futrakul et al., 2006), this paper also relied on a larger sample, thus has greater statistical power.

## Limitation

A key limitation in this study is the small number of missing values in most variables, including Apgar score. Some confidence intervals were wide, possibly due to the number of study participants, indicating lower statistical power. In addition, while some of the variables had significant effects, such as ‘Other reasons’ for admission, it may be complicated to apply within the Neotree system given this group can have a variety of admission reasons.

## Conclusion

The clinical triggers for suspecting NE and thus performing the Thompson score are not yet clearly defined. With the objective to construct an empirically-driven and parsimonious model for identifying Thompson score >10 in low-resource settings, this study identified nine risk factors that are most predictive: eight neonatal factors – Apgar score at 5 minutes, foetal heart rate at admission, hypothermia at admission, resuscitation, abnormal head shape, NE and respiratory distress as admission reasons along with others; and one intrapartum factor– maternal risk factors of sepsis other than premature rupture of membrane and offensive liquor.

The results imply that integration of these factors into the Neotree application would enable healthcare professionals to identify neonates to screen for NE using the Thompson score. However, before operationalisation into the application, further work is needed to evaluate the model’s performance through sensitivity and specificity analysis. This evaluation should ensure a large enough sample of Thompson scores to allow for the dichotomisation of predictors such as Apgar score and heart rate. Finally, future work could also focus on using more novel predictive model building techniques that use artificial intelligence and machine learning.

## Data Availability

Data is owned by the Zimbabwe Ministry of Health and cannot be made publicly available. Anonymised data is available upon request.

https://figshare.com/s/4d5f325ddf2c8ec064a3

https://doi.org/10.6084/m9.figshare.24700110

## Acknowledgements

The main research question and methods were conceived by EM in consultation with MH, FF and MCB. GC, SC, CC and LC provided feedback and suggestions for determining the *a priori* variables. NK and EC conducted the scoping review. NK prepared the data and performed data analysis with support from MCB and wrote the core manuscript. MH and MCB supported with overall feedback, editing and proofreading of the manuscript. The wider Neotree research team and staff at Sally Mugabe Central Hospital ensured data collection. This research was supported by the National Institute for Health Research (NIHR) Great Ormond Street Hospital Biomedical Research Centre. The views expressed are those of the authors and not necessarily those of the National Health Service (NHS), the NIHR or the UK Department of Health.

## Ethics

This project is a sub-study of the wider Neotree project which has research ethical approval from the Harare Central Hospital Research Ethics Committee (Reference number HCHEC070618/58) and UCL Research Ethics Committee (UCL Research Ethics Committee (reference 5019/004) including the analysis carried out in this paper. This sub-study is registered with the UCL Great Ormond Street Institute of Child Health R&D Office (R&D number 20PP42).

